# A pilot study of discovery and validation of peritoneal endometriosis biomarkers in peritoneal fluid and serum

**DOI:** 10.1101/2020.10.13.20211789

**Authors:** See Ling Loy, Jieliang Zhou, Liang Cui, Tse Yeun Tan, Tat Xin Ee, Bernard Su Min Chern, Jerry Kok Yen Chan, Yie Hou Lee

**Affiliations:** Department of Reproductive Medicine, KK Women’s and Children’s Hospital, Singapore, Singapore 229899. (SLL); (TYT); (TXE); (JKYC); Obstetrics and Gynecology-Academic Clinical Program, Duke-NUS Medical School, Singapore, Singapore 169857. (BSMC); (YHL); KK Research Centre, KK Women’s and Children’s Hospital, Singapore, Singapore 229899. (JZ); (LC); Singapore-MIT Alliance for Research and Technology, Singapore, Singapore 138602; Department of Obstetrics and Gynecology, KK Women’s and Children’s Hospital, Singapore, Singapore 229899

**Keywords:** biomarker, diagnosis, endometriosis, LC-MS/MS, metabolomics

## Abstract

**Objective:** To identify potential serum biomarkers in women with peritoneal endometriosis (PE) by first looking at its source in the peritoneal fluid (PF).

**Design:** Case-control pilot studies, comprising independent discovery and validation sets.

**Setting:** KK Women’s and Children’s Hospital, Singapore.

**Patient(s):** Women with laparoscopically confirmed PE and absence of endometriosis (control).

**Intervention(s):** None.

**Main Outcome Measure(s):** In the discovery set, we used untargeted liquid chromatography-mass spectrometry (LC-MS/MS) metabolomics, multivariable and univariable analyses to generate global metabolomic profiles of PF for endometriosis and to identify potential metabolites that could distinguish PE (n=10) from controls (n=31). Using targeted metabolomics, we validated the identified metabolites in PF and sera of cases (n=16 PE) and controls (n=19). We performed the area under the receiver-operating characteristics curve (AUC) analysis to evaluate the diagnostic performance of PE metabolites.

**Result(s):** In the discovery set, PF phosphatidylcholine (34:3) and phenylalanyl-isoleucine were significantly increased in PE than controls groups, with AUC 0.77 (95% confidence interval 0.61-0.92; *p*=0.018) and AUC 0.98 (0.95-1.02; *p*<0.001), respectively. In the validation set, phenylalanyl-isoleucine retained discriminatory performance to distinguish PE from controls in both PF (AUC 0.77; 0.61-0.92; *p*=0.006) and serum samples (AUC 0.81; 0.64-0.99; *p*=0.004).

**Conclusion(s):** Our preliminary results propose phenylalanyl-isoleucine as a potential biomarker of PE, which may be used as a minimally-invasive diagnostic biomarker of PE.

## INTRODUCTION

Endometriosis affects approximately 10% of reproductive aged women (1, 2) and is associated with substantial morbidity, including chronic pelvic pain and infertility (3, 4). Endometriosis is represented by three main subphenotypes: ovarian endometriosis (OE), superficial peritoneal endometriosis (PE) and deep infiltrating endometriosis (DIE) (5). To diagnose endometriosis, transvaginal ultrasonography can be used to detect OE and DIE (6, 7), with pelvic magnetic resonance imaging to assess the extent of DIE (1). However, the detection of PE, characterized by superficial endometrial lesions occurring on the peritoneum, remains challenging (8-11). Laparoscopic visualization remains as the standard for definitive diagnosis of PE, the most common subphenotype which accounts for ∼80% of all endometriosis (11-13).

Using laparoscopic visualization as the first line diagnostic tool poses a number of challenges, including its invasive nature, associated risks and potential complications of surgery (11). Laparoscopy is appropriate when symptoms reach a level of severity to justify the surgical risk (5), yet clinical symptoms has a poor correlation with disease burden (10). Indeed, accuracy of diagnosis is dependent on practitioners’ laparoscopic skills due to the diversity of endometriotic appearances and locations, insofar that endometriosis may be inadvertently missed with less obvious or microscopic endometriotic lesions (14). Consequently, there is often a delay with an average of eight years in the diagnosis of endometriosis (6). Thus, there is a great need to identify a less invasive method for PE diagnosis, which would have a groundbreaking impact in preventing or delaying disease progression, improving patients’ quality of life and the efficacy of available treatments. This is particularly important in women for whom fertility is a priority whereby hormonal treatment is not appropriate (13).

To date, despite the evaluation of numerous potential biomarkers, a reliable biomarker specifically for the diagnosis of PE has yet to be identified (10). Advances of high-throughput bioanalytical technologies in omics have made metabolomics a powerful tool for biomarker discovery (15, 16). Metabolites are intermediates to a wide range of biological processes and signaling axes such as mammalian target of rapamycin (mTOR), peroxisome proliferator-activated receptor (PPAR) and mitogen-activated protein kinase (MAPK) pathways (17-19). The association of aberrant metabolism and endometriosis has emerged in recent years (20-22). Differences in metabolism and metabolite levels reflecting endometriosis subphenotypes pathophysiology potentially forms the basis for identifying novel biomarkers for PE.

The peritoneal fluid (PF) is notably rich in proteins and lipids including cytokines, chemokines, growth factors and matrix metalloproteinases, and serves an important environment where endometriotic lesions reside and communicate with surrounding tissues including nerve cells and ovaries (23-25). The PF is therefore a source for the assessment of the dysregulated peritoneal cavity, and for reflecting dysregulated metabolic state of the subphenotypes. Thus, in this study, we aimed to identify potential biomarkers for the diagnosis of peritoneal endometriosis by first looking at its source in the peritoneal fluid. Through untargeted metabolomics, we characterized global metabolomics alterations in the peritoneal fluid samples, and multivariable and univariable statistics were used to discover potential biomarkers that resolve women with a laparoscopic diagnosed peritoneal endometriosis and without endometriosis. To verify the identified potential biomarkers in peritoneal fluid, we performed targeted metabolomics on peritoneal fluid and serum samples of women from independent case-control sets.

## MATERIALS AND METHODS

### Study design

In this case-control pilot study, investigation of biomarkers was conducted in two phases (Supplemental Figure 1, available online). Phase I, defined as the discovery set, untargeted metabolomics approach was employed to generate PF-specific global metabolomic maps of endometriosis. Phase II, defined as the validation set, where two separate groups of women were independently enrolled to verify the identified PF-metabolites for PE diagnosis. We performed targeted metabolomics analysis using both PF (invasive) and serum samples (minimally-invasive) among women with PE and controls. This study was conducted according to the Helsinki Declaration, and all procedures were approved by the Singhealth Centralized Institutional Review Board (reference 2010-167-D).

**Figure 1.**
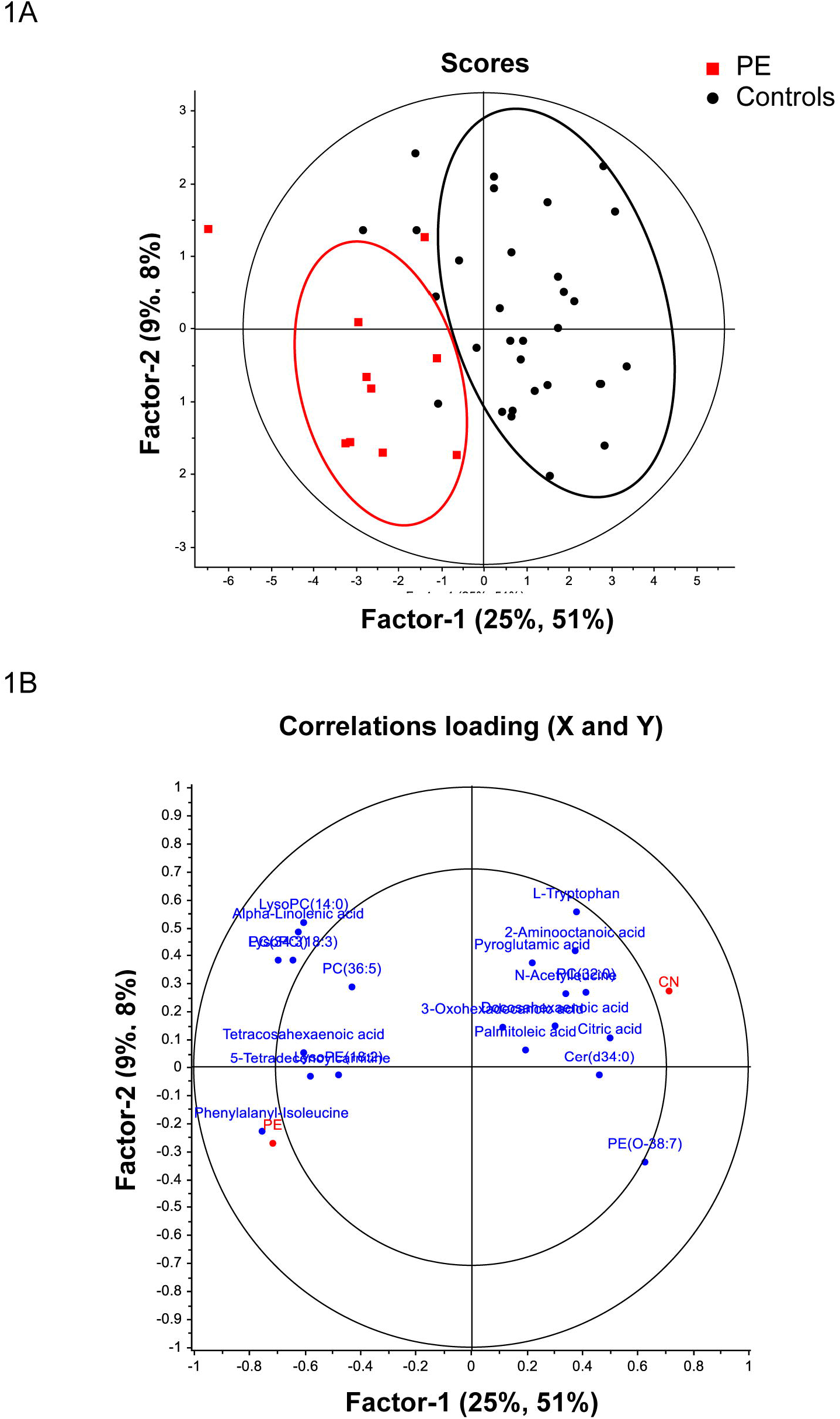
PLSR analysis of peritoneal fluid metabolites from women with PE and controls. (2A) Score plots of the discovery set samples. (2B) Loading plots of the discovery set samples. CN, control; PE, peritoneal endometriosis; PLSR, partial least squares regression.

### Clinical participants

Patients were recruited from the subfertility clinic in the KK Women’s and Children’s Hospital, Singapore, between June 2010 and May 2013 for Phase I, and between June 2010 and August 2016 for Phase II. Laparoscopy was scheduled for suspected endometriosis, infertility, sterilization procedures, and/or pelvic pain. Exclusion criteria included menstruating patients, post-menopausal patients, patients on hormonal therapy (e.g. norethisterone, combined oral contraceptive pill) for at least three months before laparoscopy, and other potentially confounding diseases such as diabetes, adenomyosis or any other chronic inflammatory diseases (rheumatoid arthritis, inflammatory bowel disease, systemic sclerosis). All eligible patients provided written informed consent upon recruitment.

During diagnostic laparoscopy, a detailed inspection of the uterus, fallopian tubes, ovaries, pouch of Douglas and the pelvic peritoneum was performed by senior gynecologists subspecialized in reproductive endocrinology and infertility. Patients with laparoscopically confirmed PE were defined as the cases, and staged according to the revised American Fertility Society classification of endometriosis (AFS, 1985; ASRM, 1997). The possible overlapping of the three lesion subphenotypes led us to classify the patients according to the worst lesion found in each subject, based on endometriosis subphenotype grouping by Somigliana et al. (26) and Chapron et al. (27). Patients from the same clinic but laparoscopically observed to be without endometriosis were defined as the controls, including those with benign gynecological presentations such as tubal occlusion, uterine fibroids, benign ovarian cysts and polycystic ovary.

### Sample collection and sample processing

For the collection of PF, clinical staff collected samples via aspiration with a syringe attached to an irrigation/suction tube from the Pouch of Douglas, with 1% protease inhibitor added (Roche, Switzerland) as previously described (28), and which is in line with Endometriosis Phenome and Biobanking Harmonization Project Standard Operating Procedures (29). The aspirates were then centrifuged (1,000×g, 4°C) for 10 min and the clear supernatants were transferred to 15 mL tubes. For the collection of peripheral venous blood, samples were collected into the BD Vacutainer® SST II tubes. After 10 min centrifugation (1,200×g, 4°C), the top yellowish layers were transferred to 15 mL tubes, followed by centrifuging for another 10 min (3,600×g, 4°C) where the supernatants were transferred to 1 ml aliquots. Both PF and serum samples were stored at −80°C until further use (28).

Prior to LC-M/S analysis, 100 μL from PF or serum sample was thawed at 4°C and proteins were precipitated with 400 μL ice-cold methanol. After vortexing for 1 min, the mixture was centrifuged at 17,000×g for 10 min at 4 °C and the supernatant was collected and evaporated to dryness in a vacuum concentrator. The dry extracts were then resuspended in 100 μL of 98:2 water/methanol or in 100 µL of 0.1% formic acid in methanol for untargeted or targeted metabolomics, respectively. Quality control (QC) samples were prepared by mixing equal amounts of reconstituted extracts from all the samples and processed as per other samples. All samples were kept at 4°C and analyzed in a random manner. QC samples are interspersed through the analytical runs and ran after each 10^th^ sample to monitor the stability of the system.

### Untargeted mass spectrometry analysis

We performed metabolomics analysis as previously described with modifications (30). Reversed-phase liquid chromatography-MS analyses were performed using the Agilent 1290 ultrahigh pressure liquid chromatography system (Waldbronn, Germany) equipped with a 6520 Q-TOF mass detector managed by a MassHunter workstation. The column used for the separation was rapid resolution HT Zorbax SB-C18 (2.1×100 mm, 1.8 μm; Agilent Technologies, USA), and the mobile phase was (A) 0.1% formic acid in water and (B) 0.1% formic acid in methanol. The initial condition of the gradient elution was set at 2% B for 2 min with a flow rate of 0.4 ml/min. A 7 min linear gradient to 70% B was then applied, followed by a 5 min gradient to 100% B which was held for 3 min. The sample injection volume was 2 μL and the oven temperature was set at 40°C. The electrospray ionization mass spectra were acquired in both positive and negative ion mode. Mass data were collected between m/z 100 and 1000 at a rate of two scans per second. The ion spray voltage was set at 4,000 V for positive mode and 3,500 V for negative mode. The heated capillary temperature was maintained at 350°C. The drying gas and nebulizer nitrogen gas flow rates were 12.0 L/min and 50 psi, respectively. Two reference masses were continuously infused to the system to allow constant mass correction during the run: m/z 121.0509 (C_5_H_4_N_4_) and m/z 922.0098 (C_18_H_18_O_6_N_3_P_3_F_24_).

### Targeted mass spectrometry analysis

The targeted LC-MS/MS analysis was performed in multiple reaction monitoring mode via Triple Quadrupole 6460 mass spectrometer with Jet Stream (Agilent Technologies). Chromatographic separation was achieved by using Eclipse Plus column C18 (2.1×50 mm; Agilent, US) with mobile phases (A) 10 mM ammonium formate and 0.1% formic acid in water and (B) 0.1% formic acid in methanol. The initial condition was set at 100% A for 3 min at a flow rate of 0.3ml/min. A 3 min linear gradient to 100% B was then applied and held for 3 min. Then the gradient returned to starting conditions over 0.1 min and maintain at the initial condition for 3 min. The column was kept at 45°C and the flow rate was 0.3 mL/min. The auto-sampler was cooled at 4°C and an injection volume of 2 μL was applied. Mass transition and collision energy were optimized for each compound by direct infusion of individual standard solutions. Both positive and negative electrospray ionization modes were performed with the following source parameters: drying gas temperature at 250°C with a flow of 5 L/min, nebulizer gas pressure at 40 psi, sheath gas temperature at 400°C with a flow of 11 L/min, capillary voltage 4,000 and 3,500 V for positive and negative mode respectively, and nozzle voltage 500 V for both positive and negative modes. Data acquisition and processing were performed using MassHunter software (Agilent Technologies, US).

### Data analysis and Compound identification

For metabolomics analysis, raw spectrometric data were converted to mzData (LC-MS/MS) and NetCDF (GC-MS) formats via Masshunter (Agilent, US) and input to open-source software MZmine 2.0 for peak finding, peak alignment and peak normalization across all samples. The structure identification of the differential metabolites was based on a previously described strategy (31). First, the element composition C_13_H_25_NO_4_ of the *m/z* 260.18 ion was calculated based on the exact mass, the nitrogen rule and the isotope pattern by Masshunter software from Agilent. Then, the elemental composition and exact mass were used for open source database searching, including LIPIDMAPS (http://www.lipidmaps.org/), HMDB (http://www.hmdb.ca/), METLIN (http://metlin.scripps.edu/) and MassBank (http://www.massband.jp/). Next, MS/MS experiments were performed to obtain structural information via the interpretation of the fragmentation pattern of the metabolite.

### Statistical analysis

We used IBM SPSS statistics, version 19 (USA), the Unscrambler and GraphPad Prism, version 7.0 (USA) for statistical analyses. For the untargeted metabolomics, we used partial least squares analysis and volcano plots to preliminarily select significant metabolites. Mann-Whitney *U*-test and Fisher’s exact test were used to compare continuous and categorical subject characteristics respectively between PE versus controls. Significantly different metabolites (SDMs) were defined by a fold change of >1.50 for increased metabolites and <0.67 for decreased metabolites. We performed the area under the receiver-operating characteristics (ROC) curve (AUC) analysis to evaluate the diagnostic performance of PE metabolites.

AUC>0.70 was defined as optimal diagnostic performance value. Sensitivity and specificity were determined at maximum Youden Index. For the discovery datasets, a two-sided *p*<0.05 was considered statistical significance. For the validation datasets, a two-sided *p*<0.025 (0.05/2 outcomes) to account for multiplicity was considered statistically significant.

## RESULTS

### Characteristics of participants

For the discovery set, PF samples were analyzed for 10 women with PE (mean age 33.3 years old) and 31 women who served as controls (mean age 33.9 years old). Majority of women with PE were at minimal-mild stage of endometriosis (rAFS stage I-II). No differences were observed in terms of age, ethnicity and cycle phase between women with PE and controls (Supplemental Table 1, available online).

For the validation set, women with PE and controls were compared and analyzed. These included PF samples for 19 PE (mean age 34.0 years old) and 20 controls (mean age 35.9 years old). Serum samples were available for 16 PE and 19 controls, as illustrated in Supplemental Figure 1 (available online). Similar to the discovery set, majority of women with PE (89.5%) were at minimal-mild stage of endometriosis. No differences in age, ethnicity and cycle phase were shown between women with PE and controls.

### Different PF metabolomic profiles between women with PE and controls

Multivariable partial least squares regression (PLSR) model was constructed to reveal the PF metabolome differences between women with PE and controls from the discovery set (Phase I). The PLSR score plots unbiasedly yielded good separation between metabolomic profiles of women with PE and controls (Figure 1A). Next, we projected the metabolomic profiles on the endometriosis subphenotypes to identify metabolites that drive the subphenotype separation. The PLSR loadings plot yielded biochemically diverse metabolites for the separation of women with PE and controls. This discrimination was primarily driven by the top 20 metabolites shown in Figure 1B. These metabolites comprised of 7 phospholipids, 5 free fatty acids, 4 amino acids, 1 carnitine, 1 sphingolipid, 1 dipeptide and 1 tricarboxylic acid cycle intermediate. The list of PF metabolites for PE and controls are shown in Supplemental Table 2 (available online). To further ascertain the relationship of key metabolites to the subphenotypes, fold change was applied to determine metabolites that are significantly different in PE compared with controls.

### Discovery of PF metabolites that distinguished PE from controls

Using volcano plots, 13 PF metabolites showed significant fold change differences (*p*<0.05) when compared PE with controls. Of these 13 metabolites, five were identified as SDMs with fold change >1.50 or <0.67. Compared with the controls, women with PE exhibited higher levels of 5-tetradecenoylcarnitine (2.33-fold), phosphatidylcholine C34:3 (1.66-fold), phenylalanyl-isoleucine (1.79-fold) and tetracosahexaenoic acid (1.72-fold), but lower level of ceramide d34:0 (0.44-fold) (Figure 2). When diagnostic performance was evaluated by ROC analysis, phenylalanyl-isoleucine and phosphatidylcholine C34:3 demonstrated diagnostic potential in distinguishing women with PE from controls, as indicated by AUC 0.98 (95% confidence interval (CI) 0.95-1.02; *p*<0.001; sensitivity 100%; specificity 95.8%) and by AUC 0.77 (95% CI 0.61-0.92; *p*=0.018; sensitivity 77.8%; specificity 71.4%), respectively (Figure 3).

**Figure 2.**
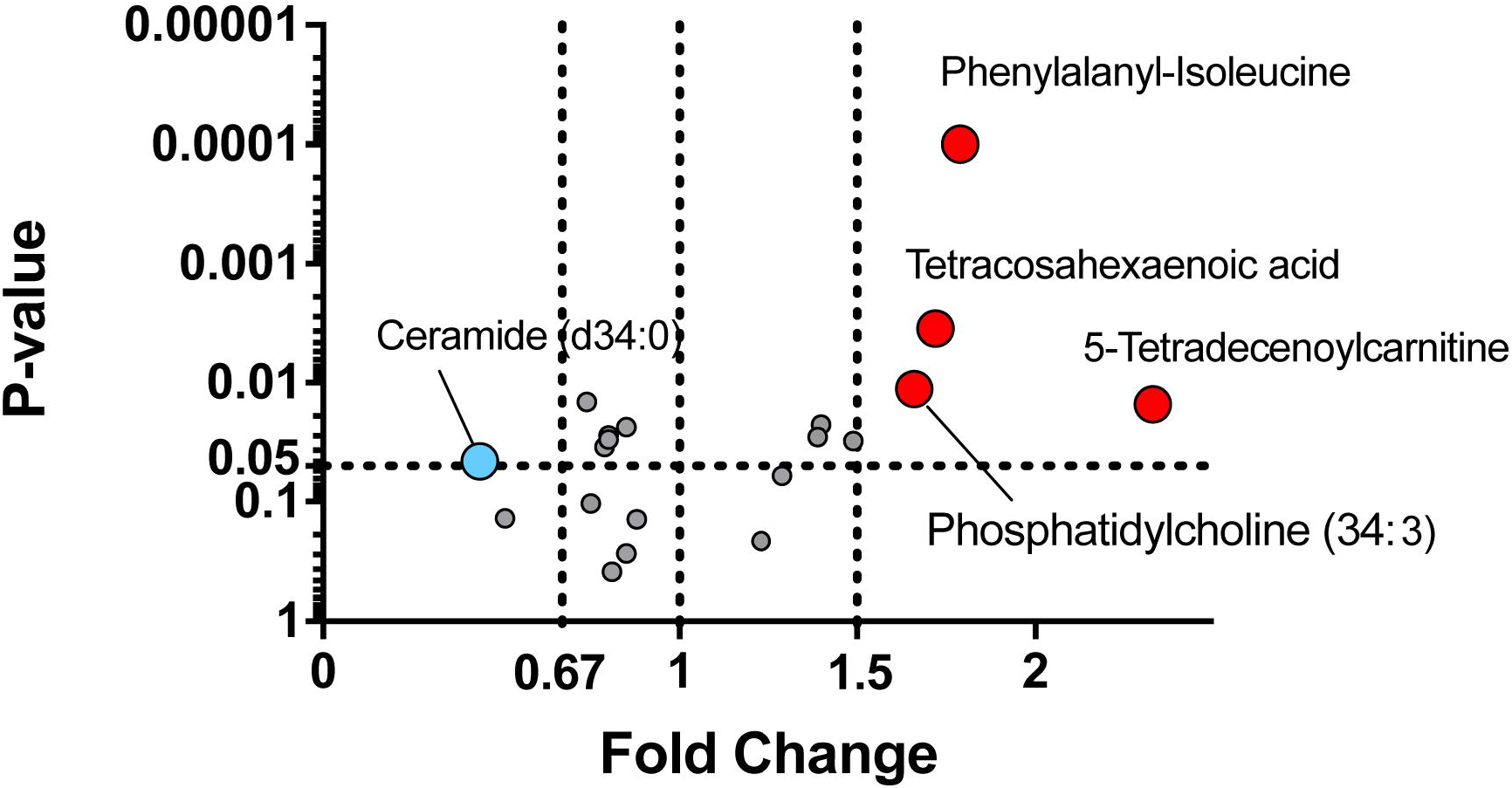
Volcano plots showing peritoneal fluid differential metabolites in women with PE relative to controls from the discovery set. Red and blue dots represent significantly different metabolites with fold change >1.5 (increased) and <0.67 (decreased), respectively (*p*<0.05). Metabolites were identified using untargeted LC-MS/MS metabolomics. LC-MS, liquid chromatography-mass spectrometry; PE, peritoneal endometriosis.

**Figure 3.**
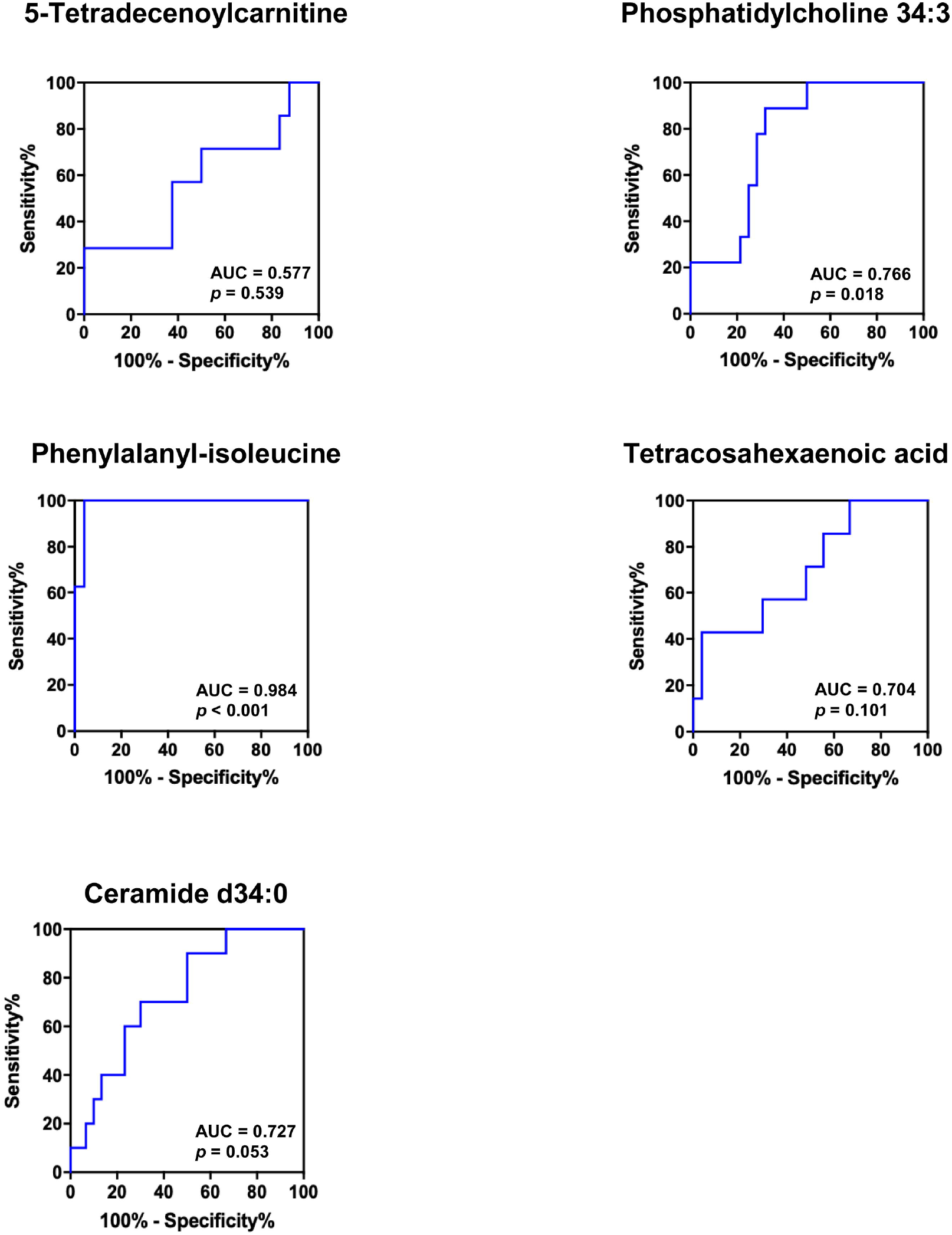
Receiver-operating characteristic curves of significantly different metabolites for peritoneal endometriosis (versus controls). Metabolites were identified using untargeted metabolomics in the discovery set. AUC of 0.5 suggests no discrimination.

### Validation of potential PF and serum metabolites that distinguished PE from controls

In Phase II, to more precisely quantify the relationship of the candidate biomarkers in PE relative to controls, we next developed a targeted metabolomics assay to validate biomarker candidates phenylalanyl-isoleucine and phosphatidylcholine C34:3. Analyzing the PF samples from an independent set of women using our developed targeted metabolomics assay, phenylalanyl-isoleucine retained its diagnostic potential in distinguishing PE from controls (AUC 0.77, 95% CI 0.61-0.92; *p*=0.006; sensitivity 73.7%; specificity 72.2%) (Figure 4A). By contrast, phosphatidylcholine C34:3 did not hold up to its initial diagnostic value (AUC 0.65, 95% CI 0.47-0.83; *p*=0.121) and hence not considered for further analysis. Next, we investigated the utility of phenylalanyl-isoleucine as a minimally-invasive biomarkers using serum samples. We found that circulating phenylalanyl-isoleucine consistently distinguished PE from controls with AUC 0.81 (95% CI 0.64-0.99; *p*=0.004; sensitivity 71.4%; specificity 100%) (Figure 4B).

**Figure 4.**
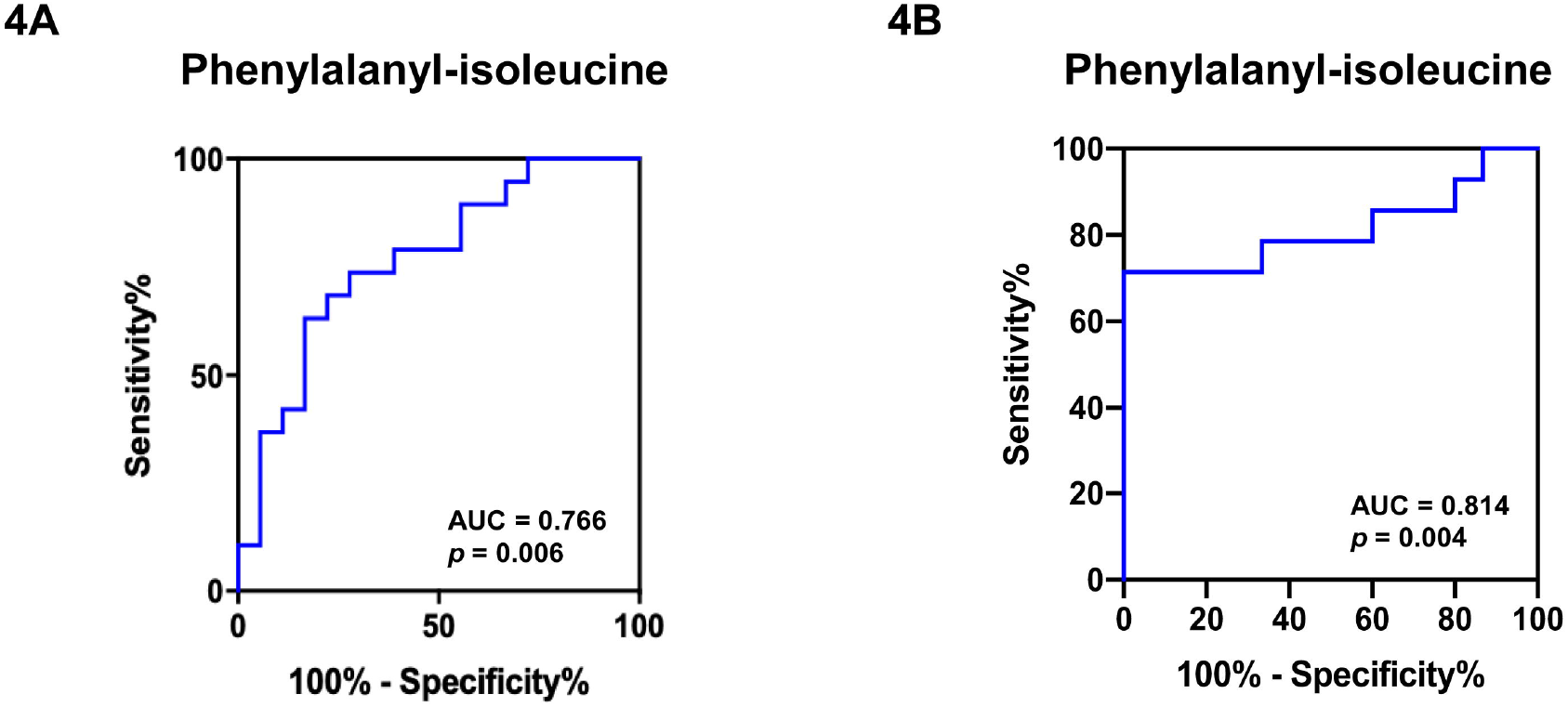
Receiver-operating characteristic curves of phenylalanyl-isoleucine for peritoneal endometriosis (versus controls) using peritoneal fluid (5A) and sera (5B). Phenylalanyl-isoleucine was identified using targeted metabolomics in the validation set. AUC of 0.5 suggests no discrimination.

## DISCUSSION

In this pilot study, we identified PF metabolites that differentiated women with PE from controls. Using untargeted metabolomic LC-MS/MS that characterized the global metabolomic alterations in the PF, a diverse PF metabolomes was shown in these women. The PF metabolomic profiles pertaining to the PE subphenotype were then linked the pathophysiological changes to the serum for biomarker discovery using a combination of untargeted and targeted metabolomics. We provided evidence showing a novel PF metabolite, phenylalanyl-isoleucine, has the potential as a biomarker of PE and was subsequently validated. Importantly, this metabolite was reflected in circulation, with a diagnostic performance value of 81.4% in serum (sensitivity 71.4%; specificity 100%), suggesting its potential use as a minimally-invasive diagnostic biomarker of PE.

PF is proximal to endometriotic lesions and thus, forms an environment that reciprocally communicates with the lesions (32, 33). This indicates that PF can be a potential useful source for biomarker discovery of PE. Intriguingly, the diagnostic characteristics of PF biomarkers for PE using metabolomics approach have not been assessed. Majority of earlier studies have focused on identifying potential biomarkers of endometriosis in general without subphenotypes specificity (21, 28, 34-37), with few focused on OE (22, 38, 39) but limited on PE (40).

Importantly, validation of proposed biomarkers which is critical to demonstrate biomarker robustness has been rarely conducted (41). In biomarker discovery studies, differential levels of metabolites such as cancer antigen 125 (CA-125), carnitines, phosphatidylcholine and sphingomyelin in PF were observed in women with endometriosis (28, 38, 42), but there is seldom linkage of these biomarkers to the circulating levels. CA-125 is a commonly investigated biomarker for endometriosis despite its undefined role in primary diagnosis (3, 10). As suggested, we performed additional analysis for CA-125 in serum of women from the validation set and compared with its performance with serum phenylalanyl-isoleucine. Serum CA-125 showed poor diagnostic performance (AUC 0.66; 95% CI 0.47-0.84; *p*=0.120) in differentiating PE (n=19) from controls (n=16) (Supplemental Figure 2, available online). Our findings are consistent with international guidelines, such as the National Institute for Health and Care Excellence (NICE) and the European Society of Human Reproduction and Embryology (ESHRE), which have made recommendations to not use serum CA-125 as biomarker for endometriosis diagnosis due to its limited diagnostic performance (13, 43).

Our results demonstrate that phenylalanyl-isoleucine was increased in PF and serum of PE women, with a high discriminatory ability to distinguish between women with PE and other gynecological disorders requiring laparoscopic diagnosis (controls). Thus, data obtained from this study have connected pathophysiology in the dysregulated peritoneal cavity to the circulation in endometriosis, in line with reports showing correlations between biomarkers in PF and serum (44, 45). Phenylalanyl-isoleucine (C_15_H_22_N_2_O_3_) is a dipeptide composed of phenylalanine and isoleucine (46), which has been shown to play an essential role in intracellular signal transduction (47). However, its functional role in endometriosis pathophysiology remains unclear. To the best of our knowledge, phenylalanyl-isoleucine is a novel metabolite which has not been reported for endometriosis. Increased phenylalanyl-isoleucine in PF might indicate an altered metabolic state that would potentially contribute to further growth and development of peritoneal lesions (48). Compared to healthy controls, women with early stage of endometriosis had previously found to exhibit decreased phenylalanine and isoleucine in endometrial tissue and serum, respectively (20, 37). In contrast, phenylalanyl-isoleucine appeared to be relatively abundant in PF of women with PE, which could be explained by the proximity of PF with endometriotic lesions implanted in the peritoneal cavity. It is possible that increased phenylalanyl-isoleucine reflected shedding into the PF due to high levels of cell division, cell death and protein degradation. This is supported by Li et al. (49) which found upregulations of various amino acids in the eutopic endometrium of women with early endometriosis. Further research is required to elucidate the association of phenylalanyl-isoleucine with PE pathophysiology.

Several studies have classified women with endometriosis according to their disease stage, where greater altered metabolomic profiles and metabolite levels were seen in women with later stages (III-IV) of endometriosis as compared with the controls (44, 45). Importantly, differences in the levels and types of metabolites were also shown for women with stage I and II endometriosis in relation to the controls (37, 49), indicating their potential for early disease diagnosis and prognosis. In this study, PE cases were mostly classified at minimal-mild stages (stage I-II), suggesting the candidate metabolite in women with PE might be additionally useful to reflect the early stage of the disease.

Strengths of this study included quantitative and structured methodological framework to identify the potential biomarker of PE, with the applications of untargeted, unbiased metabolomics in the discovery process and targeted metabolomics with exquisite assay sensitivity and precision in the validation process. Importantly, this study captured the most comprehensive catalogued metabolome space of PF in women with laparoscopic diagnosed PE to-date. By treating the identification of PF biomarker as a pre-selection procedure for subsequent study for serum marker, we successfully demonstrated the presence of candidate biomarker in sera of PE women from an independent group, with optimal diagnostic performance for PE. The procedure of validating biomarker using separate, independent sets of samples by applying same analytical approach has rarely been carried out in most previous studies. However, an important limitation of this study was the small sample size. This has restricted our capability to stratify women with PE into individual stage of disease for further assessing the prognostic applications of phenylalanyl-isoleucine. However, preliminary findings generated from this pilot study serves as important baseline supportive evidence for conducting subsequent larger scale studies to confirm the reliability and validity of phenylalanyl-isoleucine for PE diagnosis.

## CONCLUSIONS

In conclusion, we have identified a signature metabolite known as phenylalanyl-isoleucine in PF of women with PE through untargeted and targeted metabolomics in independent datasets, suggesting its involvement in the pathophysiology of PE. The same metabolite was identified in the sera of women with PE. Large scale metabolomics studies validating this biomarker across different populations will further determine its diagnostic robustness. More studies are also warranted to test for the robustness of this biomarker in distinguishing PE from other subphenotypes of endometriosis and pelvic inflammatory diseases that exhibit similar clinical symptoms.

## Data Availability

Raw data are available from the corresponding author.

## Acknowledgements

We thank the participants of the study. This work was supported by the National Research Foundation Singapore under its National Medical Research Council Centre Grant Program (NMRC/CG/M003/2017) and administered by the Singapore Ministry of Health’s National Medical Research Council, and Duke-NUS Office of Academic Medicine (Clinical and Translational Research in Endometriosis). JCKY received salary support from Singapore’s Ministry of Health’s National Medical Research Council (CSA-SI-008-2016).

## Supplementary data

**Supplemental Figure 1**. Flow diagram illustrating the study design. Phase II comprised of an independent set of patients from phase I which was used to validate the PE metabolites. LC-MS/MS, liquid chromatography-mass spectrometry; PE, peritoneal endometriosis; ROC, receiver operating characteristics; SDMs, significantly different metabolites

**Supplemental Figure 2**. Receiver-operating characteristic curves of serum cancer antigen 125 (CA-125) for peritoneal endometriosis (versus controls) in women from validation dataset.

**Supplemental Table 1**. Participants’ characteristics according to types of endometriosis in the discovery and validation sets.

**Supplemental Table 2**. Peritoneal fluid metabolites of women with peritoneal endometriosis and controls, identified using untargeted metabolomics analysis in the discovery set.

