## Supplementary materials for "A pilot study of discovery and validation of peritoneal endometriosis biomarkers in peritoneal fluid and serum"

**Phase II**

**Phase I**

**Validation set**

**Discovery set**

**Serum**

- 16 PE
- 19 Controls

**Peritoneal fluid**

- 19 PE
- 20 Controls

**Peritoneal fluid**

- 10 PE
- 31 controls

Untargeted LC-MS/MS metabolomics

Targeted LC-MS/MS metabolomics

**PE vs. Controls**

**-**

**PE vs. Controls**

**PE vs. Controls**

Multivariate & Univariate statistical analyses

Discovery of PE metabolites

ROC curve analysis

ROC curve analysis

SDMs

SDMs

Validation of potential PE biomarkers

SDMs

ROC curve analysis

**Supplemental Figure 1.** Flow diagram illustrating the study design. Phase II comprised of an independent set of patients from phase I which was used to validate the PE metabolites. LC-MS/MS, liquid chromatography-mass spectrometry; PE, peritoneal endometriosis; ROC, receiver operating characteristics; SDMs, significantly different metabolites

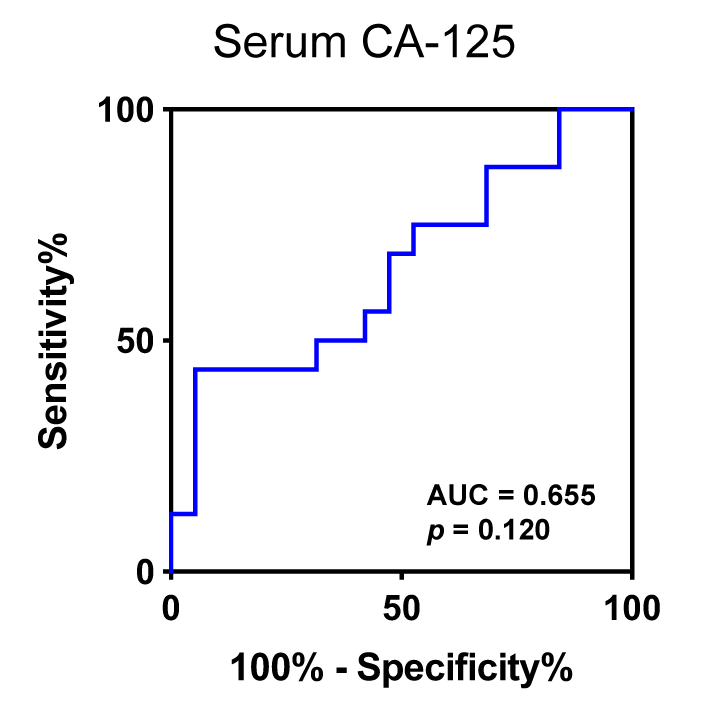

**Supplemental Figure 2.** Receiver-operating characteristic curves of serum cancer antigen 125 (CA-125) for peritoneal endometriosis (versus controls) in women from validation dataset.

**Supplemental Table 1.** Participants’ characteristics according to types of endometriosis in the discovery and validation sets.

|  | Discovery set | | |  | Validation set | | |
| --- | --- | --- | --- | --- | --- | --- | --- |
| Characteristics | PE | Controls | PE vs. Controls |  | PE | Controls | PE vs. Controls |
|  | (*n*=10) | (*n*=31) | *p*-value^a^ |  | (*n*=19) | (*n*=20) | *p*-value^a^ |
| Endometriosis stage, n (%) |  |  |  |  |  |  | - |
| Stage I & II | 8 (80.0) | - | - |  | 17 (89.5) | - |  |
| Stage III & IV | 2 (20.0) | - | - |  | 2 (10.5) | - |  |
| Age (years) | 33.3 ± 4.1 | 33.9 ± 7.1 | 0.790 |  | 34.0 ± 4.1 | 35.9 ± 5.6 | 0.236 |
| Ethnicity, n (%) |  |  | 0.783 |  |  |  | 0.358 |
| Chinese | 7 (70.0) | 20 |  |  | 12 (63.2) | 14 (70.0) |  |
| Malay | 2 (20.0) | 5 |  |  | 5 (26.3) | 2 (10.0) |  |
| Others^b^ | 1 (10.0) | 6 |  |  | 2 (10.5) | 4 (20.0) |  |
| Cycle phase, n (%) |  |  | 0.836 |  |  |  | >0.995 |
| Proliferative | 3 (30.0) | 12 (38.7) |  |  | 10 (52.6) | 10 (50.0) |  |
| Secretory | 7 (70.0) | 17 (54.8) |  |  | 9 (47.4) | 10 (50.0) |  |
| Menopause | 0 | 0 |  |  | 0 | 0 |  |
| Menses | 0 | 0 |  |  | 0 | 0 |  |
| Unknown | 0 | 2 (6.5) |  |  | 0 | 0 |  |

Values are means ± standard deviations) or n (%). PE, peritoneal endometriosis.

^a^Based on independent t-test for continuous variables or Fisher’s exact test for categorical variables.

^b^Other ethnicities comprised of Burmese, Indians, Indonesians, Filipinos and Vietnamese.

**Supplemental Table 2.** Peritoneal fluid metabolites of women with peritoneal endometriosis and controls, identified using untargeted metabolomics analysis in the discovery set.

|  |  | Controls | PE |  | PE vs. Controls | |
| --- | --- | --- | --- | --- | --- | --- |
| No. | Peritoneal fluid metabolites | Average intensity | |  | Fold change | *p*-value |
| 1 | 2-Aminooctanoic acid | 57486 | 45905 |  | 0.80 | 0.028 |
| 2 | 3-Oxohexadecanoic acid | 370315 | 315328 |  | 0.85 | 0.271 |
| 3 | 5-Tetradecenoylcarnitine | 207944 | 485453 |  | **2.33** | **0.015** |
| 4 | Alpha-linolenic acid | 703713 | 983484 |  | 1.40 | 0.023 |
| 5 | Ceramide (d34:0) | 312041 | 136905 |  | **0.44** | **0.046** |
| 6 | Citric acid | 349602 | 276879 |  | 0.79 | 0.035 |
| 7 | Docosahexaenoic acid | 2203865 | 1654924 |  | 0.75 | 0.103 |
| 8 | L-Tryptophan | 163957 | 139566 |  | 0.85 | 0.024 |
| 9 | LysoPC(14:0) | 5618828 | 7271356 |  | 1.29 | 0.060 |
| 10 | LysoPC(18:3) | 2253623 | 3124839 |  | 1.39 | 0.029 |
| 11 | LysoPE(18:2) | 992887 | 1477665 |  | 1.49 | 0.031 |
| 12 | N-Acetylleucine | 743384 | 381151 |  | 0.51 | 0.138 |
| 13 | Palmitoleic acid | 1927818 | 1568409 |  | 0.81 | 0.388 |
| 14 | PC(32:0) | 10443082 | 8310943 |  | 0.80 | 0.030 |
| 15 | PC(34:3) | 3077537 | 5101979 |  | **1.66** | **0.011** |
| 16 | PC(36:5) | 643302 | 792676 |  | 1.23 | 0.213 |
| 17 | PE(O-38:7) | 2522637 | 1868547 |  | 0.74 | 0.014 |
| 18 | Phenylalanyl-isoleucine | 166115 | 297664 |  | **1.79** | **<0.001** |
| 19 | Pyroglutamic acid | 977158 | 856031 |  | 0.88 | 0.140 |
| 20 | Tetracosahexaenoic acid | 746077 | 1282879 |  | **1.72** | **0.003** |

Significantly different metabolite with fold change >1.5 for increased metabolite or <0.67 for decreased metabolite are represented in bold, *p*<0.05. PE, peritoneal endometriosis; PC, Phosphatidylcholine; PE, phosphatidylethanolamine.
